# Shared within-host SARS-CoV-2 variation in households

**DOI:** 10.1101/2022.05.26.22275279

**Authors:** Katharine S. Walter, Eugene Kim, Renu Verma, Jonathan Altamirano, Sean Leary, Yuan J. Carrington, Prasanna Jagannathan, Upinder Singh, Marisa Holubar, Aruna Subramanian, Chaitan Khosla, Yvonne Maldonado, Jason R. Andrews

## Abstract

**Background:** The limited variation observed among SARS-CoV-2 consensus sequences makes it difficult to reconstruct transmission linkages in outbreak settings. Previous studies have recovered variation within individual SARS-CoV-2 infections but have not yet measured the informativeness of within-host variation for transmission inference.

**Methods:** We performed tiled amplicon sequencing on 307 SARS-CoV-2 samples from four prospective studies and combined sequence data with household membership data, a proxy for transmission linkage.

**Results:** Consensus sequences from households had limited diversity (mean pairwise distance, 3.06 SNPs; range, 0-40). Most (83.1%, 255/307) samples harbored at least one intrahost single nucleotide variant (iSNV; median: 117; IQR: 17-208), when applying a liberal minor allele frequency of 0.5% and prior to filtering. A mean of 15.4% of within-host iSNVs were recovered one day later. Pairs in the same household shared significantly more iSNVs (mean: 1.20 iSNVs; 95% CI: 1.02-1.39) than did pairs in different households infected with the same viral clade (mean: 0.31 iSNVs; 95% CI: 0.28-0.34), a signal that increases with increasingly liberal thresholds.

**Conclusions:** Although only a subset of within-host variation is consistently shared across likely transmission pairs, shared iSNVs may augment the information in consensus sequences for predicting transmission linkages.

## Background

SARS-CoV-2 genomic sequencing has been powerfully used to reconstruct the virus’ evolutionary dynamics at broad temporal and spatial scales[1–3]. Yet the virus’ relatively slow substitution rate compared with its short serial interval limits the viral diversity observed in many outbreaks, and viral consensus sequences—which represent the most common allele along the viral genome—are often identical or nearly so [4,5].

In superspreading events, identical consensus sequences have provided important evidence of recent shared transmission. For example, four individuals on the same international flight were infected with identical SARS-CoV-2 consensus genomes, evidence that the virus could be transmitted during air travel[6]. Genomic surveillance in Boston during 2020 reported that 59 out of 83 (71%) genomes sequenced from a skilled nursing facility were identical, implicating transmission within the facility[7]. Similarly, 75% of SARS-CoV-2 consensus sequences from a fishing boat outbreak were identical to at least one other sequence, and the remaining sequences were closely related, suggesting rapid transmission from a single viral introduction [8].

While consensus sequences have been harnessed to implicate or exclude the possibility of a shared recent transmission history, closely related consensus sequences are often not sufficient for reconstructing transmission linkages. For example, in hospital-based surveillance in Wisconsin, many healthcare workers implicated in different epidemiological clusters were infected with identical SARS-CoV-2 genomes[9]. In a hospital outbreak in Portugal, groups of identical consensus sequences shared between healthcare workers and patients were frequently identified[5]. Similarly, in hospital-based surveillance in the UK, 159 of 299 (53%) genomes sampled from one hospital were identical to at least one other sampled genome[10]. While many pairs of individuals infected with identical genomes had strong or intermediate evidence of transmission, 22% had no epidemiological evidence of transmission[10], potentially the result of incomplete epidemiological information (cryptic transmission) or limited genomic variation resulting in identical, epidemiologically unlinked consensus genomes. In the absence of detailed epidemiological data, such as contact information or spatial information that might be available in hospital-based studies, it is not yet known whether routine sequencing data alone can be used to reconstruct transmission linkages of who-infected-whom or identify locations or individuals that may drive transmission.

Genomic studies of HIV and other viral and bacterial pathogens have begun to harness the pathogen variation within individual infections, or within-host diversity, to reconstruct transmission linkages [11–13]. Previous studies have reported low levels of SARS-CoV-2 diversity within individual hosts and have estimated the size of a narrow transmission bottleneck which limits the viral diversity shared across hosts[8,9,14,15]. However, more research is needed to quantify the informativeness of within-host SARS-CoV-2 variation and evaluate the effects of variant identification approaches on transmission inferences[15].

To investigate the potential for within-host SARS-CoV-2 diversity to be harnessed for studies of transmission, we deep sequenced SARS-CoV-2 samples collected from household members, allowing us to directly compare shared within-host variants among epidemiologically linked individuals and those with no known linkage, providing a test case for the transmission information contained within individual infections. We additionally sequenced artificial mixtures of SARS-CoV-2 variants to examine tradeoffs between sensitivity and specificity in within-host variation identification.

## Methods

### Collection of residual SARS-CoV-2 samples for deep sequencing

We assembled a collection of samples from four prospective SARS-CoV-2 research studies: (a) a prospective household transmission study, in which index cases with at least one reverse transcription quantitative polymerase chain reaction-confirmed (RT-qPCR) SARS-CoV-2 test were enrolled along with household members. Participants were tested daily for SARS-CoV-2 RNA via RT-qPCR, using self-collected lower nasal swabs, and households were followed until all members tested negative for seven consecutive days[16]. (b) A randomized, single-blind, placebo-controlled trial of Peginterferon Lambda-1a (Lambda) for reducing the duration of viral shedding or symptoms[17] in which oropharyngeal swabs were collected for 28 days following enrollment. (c) A phase 2 double-blind randomized controlled outpatient trial of the antiviral favipiravir for reducing the duration of viral shedding in which participants self-collected daily anterior nasal swabs for 28 days following enrollment[18]. Neither Lambda nor favipiravir was found to shorten the duration of SARS-CoV-2 viral shedding[17,18]. (d) A study of a noninvasive mask sampling method to quantify SARS-CoV-2 shedding in exhaled breath[19].

All study participants provided written consent and all studies were approved by the Institutional Review Board of Stanford University (Numbers: 55479, 57686, 56032, and 55619). We identified household members through participation in the household transmission study and address matching.

### Sample RT-qPCR testing

We collected nasal swabs in 500 µL of Primestore MTM (Longhorn Vaccines & Diagnostics) RNA-stabilizing media. For exhaled breath samples, we extracted RNA from gelatin membrane filters processed in 1-mL PrimeStore MTM media. RNA was extracted using the MagMAX Viral/Pathogen Ultra Nucleic Acid Isolation Kit (Cat # A42356 Applied Biosystems) and eluted in 50 µL of elution buffer [15] (Supplementary Methods).

### Library preparation and sequencing

We followed the ARTIC v3 Illumina library preparation and sequencing protocols[20] and sequenced amplicons on an Illumina MiSeq platform (Supplementary Methods). Sequence data is available at SRA (BioProject ID: PRJNA842503).

### Variant identification

We used the *nf-core/viralrecon v*.*2*.*4* bioinformatic pipeline to perform variant calling and generate consensus sequences from raw sequencing reads[21]. Briefly, we aligned reads to the MN908947.3 reference genome with *Bowtie 2*[22], removed primer sequences with *iVar*[23], and called variants with respect to the reference genome with *iVar*[23]. We also used this pipeline to remove reads mapping to the host genome with *Kraken2*[24], map reads with *Bowtie 2*[22], generate consensus sequences with *bcftools*[25], and assign Nextclade lineages[26]. We modified the pipeline to include variants with an alternate allele frequency ≥ 0.2%.

We included samples with a median coverage of 100X and with >70% of the genome covered by a depth of >10X. We focused our analysis on single nucleotide polymorphism (SNP) variants and excluded SNPs occurring at previously reported problematic sites[27].

To test whether commonly applied filters would improve overall accuracy, we applied five variant filters: a filter for iSNV quality from *iVar*[23] (PASS = TRUE), a variant quality score filter (Phred score >40), a depth filter (of both major and minor alleles > 5X), a filter of false positive iSNVs repeated in more than one sample in the artificial strain mixture experiment (below), and all filters. We additionally excluded iSNVs occurring in primer binding sites (except for the unfiltered variant set).

To identify shared within-host diversity across samples, we compared each unique pair of samples meeting our quality criteria. We identified shared iSNVs as shared variant positions in which a variant was not fixed and with the same alternate allele call. We additionally determined the geometric mean of the sum of minor allele frequencies at shared iSNVs for each sample in a pair as a measure of shared viral population diversity. To exclude potential shared iSNVs attributable to sequencing batch, we excluded samples sequenced on the same Illumina sequencing lane in pairwise comparisons.

### Statistical analysis

We fit a Poisson regression model for the number of iSNVs identified within a single sample including sequencing batch and participant as random effects. We additionally fit a Poisson regression model for the number of pairwise shared iSNVs as a function of pair type and distance between consensus sequences, including pair as a random effect. Finally, we fit a binomial regression model for predicting household membership as a function of the number of shared pairwise iSNVs and an indicator variable for close consensus sequences (pairwise distance ≤ 1 SNP), including the earliest samples collected from each pair to exclude multiple pairwise comparisons. We fit all models with the R package *lme4*[31], and included the set of variants after applying all filters, including iSNVs with a minor allele frequency of ≥0.2%. We excluded samples sequenced in the same sequencing batch.

### Replicating analysis in an independent deep sequencing dataset from Wisconsin

We additionally investigated patterns of shared within-host variation in a previously published dataset from a household transmission study in Wisconsin[9]. Specifically, we re-analyzed variants called by the previous study and filtered to include iSNVs with a minor allele frequency ≥1% and to exclude variants occurring at primer binding sites[9].

Samples in the previous study were sequenced in duplicate. To generate a set of variant calls that were comparable to those from our California dataset, we took the union of iSNVs identified in each replicate sample; for iSNVs detected in both samples, we included the iSNV with the greater minor allele frequency. As in the previous study, we excluded iSNVs called in genomic positions <54 or >29,837 or at position 6669, which was identified as a problematic site. As above, we excluded positions previously reported as problematic sites[27] from variant calls.

## Results

### Assembling a collection of longitudinally sampled individuals and transmission pairs

We aggregated residual nasal swabs from four studies collected from March 2020 through May 2021 and deep sequenced SARS-CoV-2 genomes using the ARTIC v3 tiled amplicon sequencing protocol (Fig. 2a). 307 SARS-CoV-2 sequences from 286 unique biological samples from 135 participants met our quality and coverage filters, including 130 samples from 32 individuals in 14 households and 57 longitudinally sampled individuals.

Samples had a median coverage depth of 1714 reads with a median of 99.0% of the SARS-CoV-2 genome covered by at least 10 reads. As expected, coverage depth was inversely correlated with RT-qPCR cycle threshold (Pearson’s r = -0.15, p=0.012), reflecting a positive correlation with SARS-CoV-2 burden.

Samples were distributed across many of the major SARS-CoV-2 lineages circulating at the time of collection (Fig. 2b). Overall, consensus sequences had a mean pairwise distance of 37.4 fixed SNPs (range, 0-76) (Fig. 2b). 193 of 456 pairs of consensus sequences (42.3%) sampled longitudinally from the same individual differed by 0-1 SNP (mean pairwise distance, 2.37; range, 0-22). The single individual with consensus sequences that differed by 22 SNPs had been a participant in a SARS-CoV-2 clinical trial and had received the antiviral drug favipiravir[18]; samples were taken 10 days apart. 251 of 778 (32.3%) of pairs of consensus sequences sampled from different individuals in the same household differed by 0-1 SNP (mean pairwise distance, 3.06; range, 0-40), consistent with the relatively slow SARS-CoV-2 substitution rate[4]. In contrast, only a small minority, 20 of 41,053 (0.49%) pairs of consensus sequences sampled from different households were within 0-1 SNP (mean pairwise distance, 38.52; range 0-76).

### A subset of within-host diversity is consistently recovered over time

A major challenge in studies of within-host pathogen diversity is in distinguishing true, low frequency intrahost nucleotide variants (iSNVs) from sequencing or bioinformatic errors[32]. By sequencing artificial strain mixtures of the Alpha and Beta variants, we established that we could reliably recover minority variants to minor allele frequencies as low as 0.25% with 10^3^ viral copies/mL (Fig. 1; Supplementary Methods; Supplementary Text), with a minimal cost of false positive iSNVs (Fig. S1).

**Figure 1.**
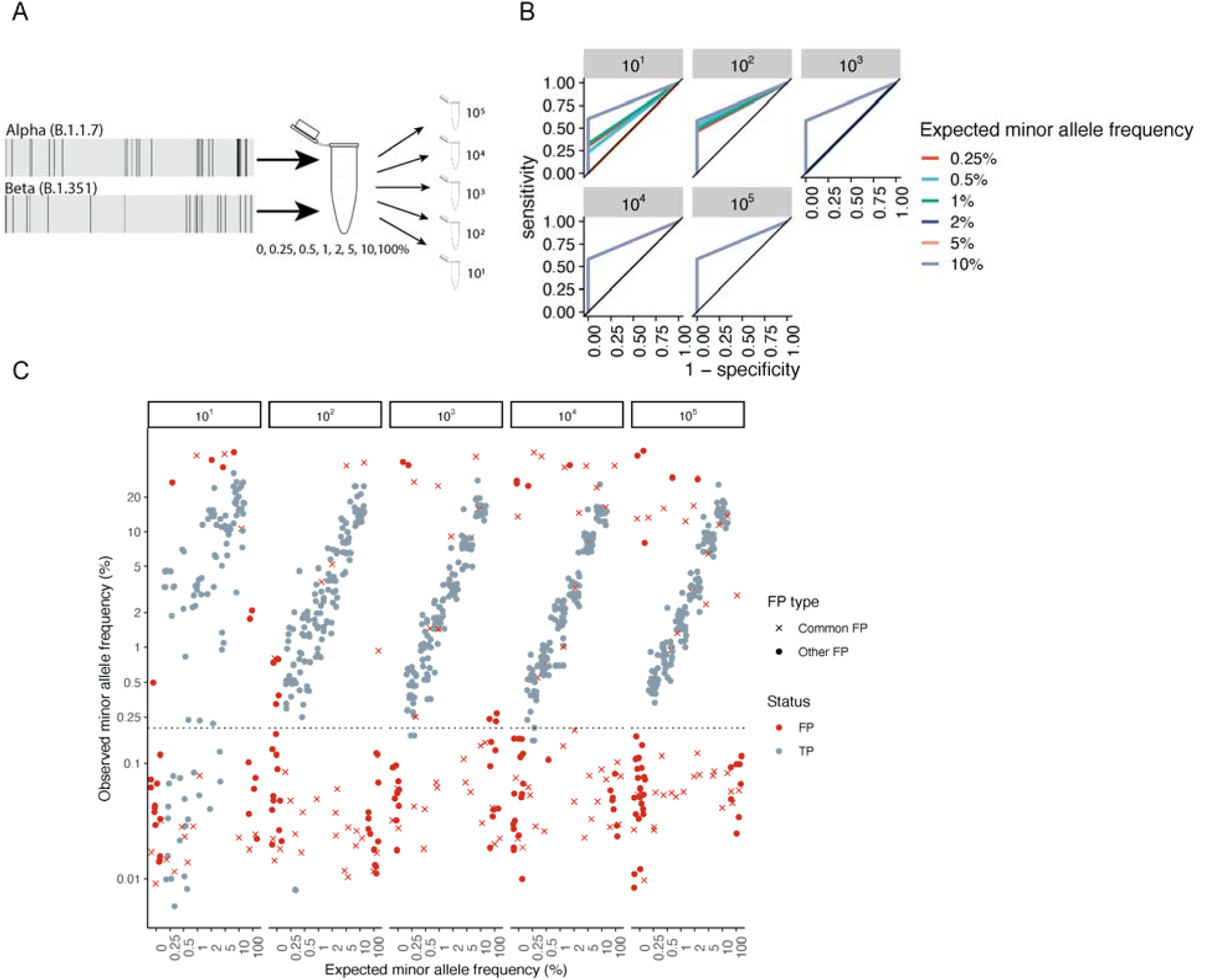
Measuring the accuracy of within-host SARS-CoV-2 variant identification. (a) Diagram of artificial strain mixture experiment. We conducted a serial dilution experiment, mixing synthetic RNA controls (Twist Biosciences) of the Alpha (B.1.1.7) and Beta (B.1.351) variants so that the minor variant comprised 0-10% of the source material and then serially diluted mixtures to a total of 10^1^-10^5^ total RNA copies. We conducted amplicon-based sequencing of artificial mixtures, identified iSNVs with the viralrecon pipeline[21], and determined the sensitivity and specificity of our variant calling pipeline to recovering true variation within our synthetic mixtures. (b) Receiver operator characteristic curve showing 1-specificity versus sensitivity in the recovery of true minority variants, colored by total RNA dilution. Lines corresponding to each dilution include results from five artificial strain mixtures, including minority variants present at 0.5-10% of the total viral pool. (c) Observed minor allele frequency versus expected minor allele frequency for artificial strain mixtures. Points indicate iSNV assignment, (FP: false positive iSNV; TP; true positive iSNV). For FP iSNVs, point shape indicates whether FPs were commonly repeated across samples (Common FP: FP identified in 10 or more samples; Other FP: any other FP iSNV). Points were jittered for visualization. The grey dotted line indicates the minor allele frequency threshold of 0.2%, below which the majority of FP iSNVs occur. Horizontal facets indicate the synthetic RNA copy number in units of genome copies per microliter.

**Figure 2.**
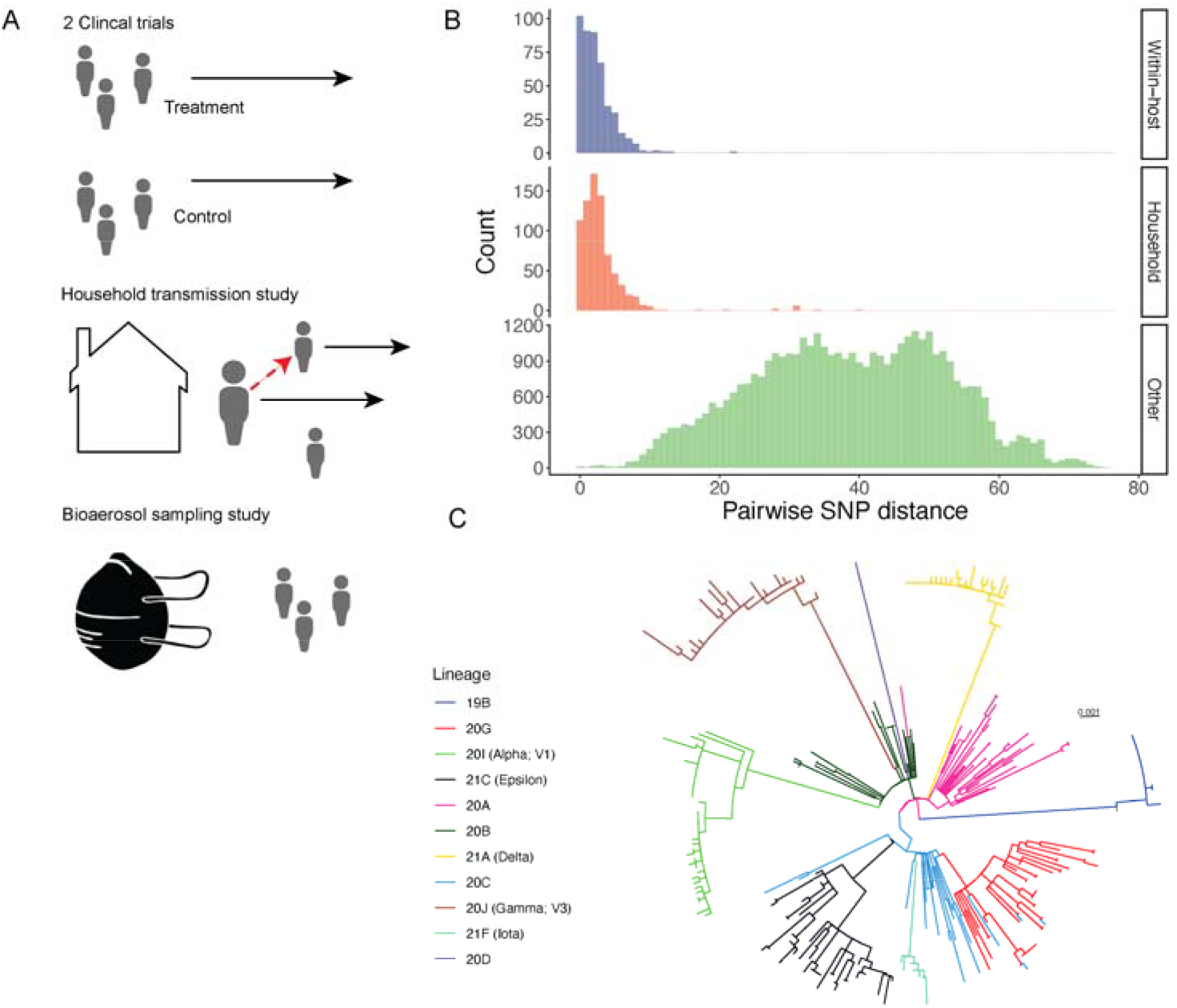
Genetic diversity of sampled SARS-CoV-2. (a) We identified household transmission pairs and longitudinal samples by address matching from four participants enrolled in studies including a household transmission study, clinical trials of Favipiravir and Lambda, and a mask shedding study. (b) Histogram of pairwise single nucleotide polymorphism (SNP) distances between consensus sequences from samples longitudinally sampled from the same individual, from different individuals within the same household, and between individuals from different households. (c) A maximum likelihood phylogeny inferred from consensus sequences with IQ-Tree with branches colored by clade. Clade assignments are made with Nextclade[43] through the nf-core viralrecon pipeline[44]. Branch lengths are in distances of substitutions per site.

Most (83.1%, 255/307) samples harbored at least one iSNV (median: 7; IQR: 2-20) above a minor allele frequency of 1.0% and applying all filters. As expected, the magnitude of recovered within-host diversity increases to a median of 20 iSNVs (IQR:4-44) and 27 (IQR:6-55) with more liberal minor allele frequency thresholds of 0.5% and 0.2% respectively (Fig. S2).

As previously reported[9,33,34], iSNVs are not consistently recovered within serial samples. Among individuals with recovered within-host diversity, a mean of 8.7% within-host iSNVs above a minor allele frequency of 1.0% and applying all filters were recovered one day later; this proportion declined with time between samples, though not significantly (r = -0.11, p = 0.23). When including unfiltered iSNVs above a 0.5% threshold, a mean of 15.4% of within-host iSNVs were recovered one day later. The variable recovery of within-host variation is consistent with previous reports that minor allele frequencies are poorly correlated within longitudinally-sampled individuals[35], potentially reflecting both sampling or sequencing bottlenecks as well as a dynamic within-host viral population (Box 1).

Despite this, the pool of minority variants recovered within an individual is a consistent marker of individual host. Pairs of sequences serially sampled from the same host consistently share 8.83 times more iSNVs (mean 0.35 shared iSNVs: 95% CI: 0.22-0.48) compared to pairs of samples from different individuals (mean 0.040 shared iSNVs: 95% CI: 0.037-0.042), after filtering and excluding samples sequenced on the same batch (Figs. 3, S2, S3). The host-specific signature declines as an increasingly strict minor allele frequency threshold is applied.

**Figure 3.**
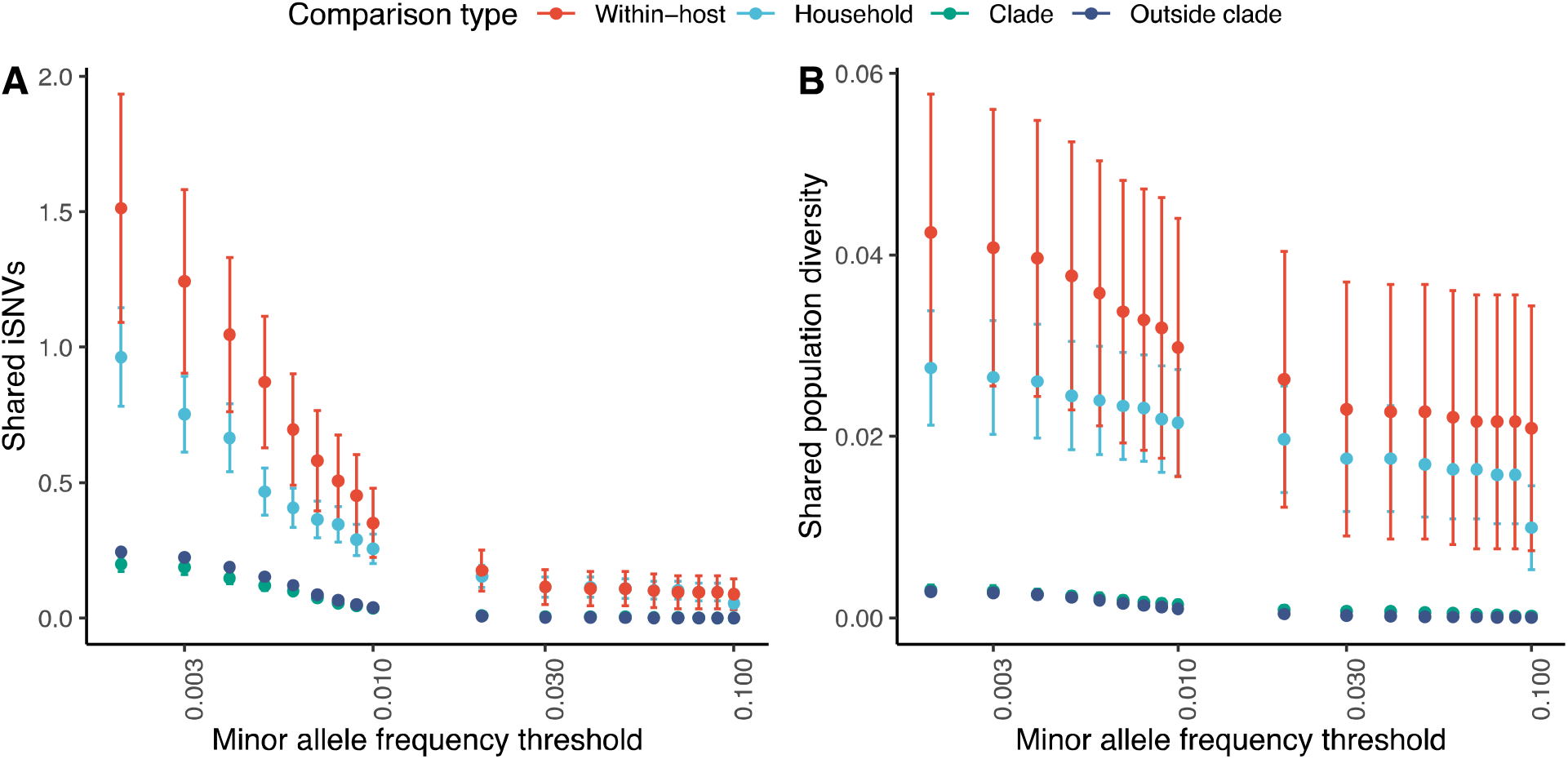
Shared within-host variants hold a signal of SARS-CoV-2 transmission. For our genomic collection from California, (a) pairwise comparisons of the number of shared iSNVs, defined as a shared minor allele present at the same genomic position, identified across different minor allele frequency thresholds, after applying all variant filters (Methods). Points and error bars indicate mean and 95% confidence intervals and are colored by comparison type. Each pair is assigned to a unique category. Within-host: pairs of samples from the same individual collected on different days; Household: pairs of individuals from the same household; Clade: pairs of individuals outside households infected with the same Nextclade clade; and Outside clade: pairs of individuals outside households infected with different Nextclade clades. Pairwise comparisons include only samples sequenced in different sequencing batches.

Within-host diversity, as measured by sample iSNV richness, was positively associated with PCR Ct value, a measure of viral burden (aOR: 1.08; 95% CI: 1.08-1.10), in a general linear model, including batch and participant as random effects. Among samples with symptom information, days following symptom onset was not associated with increased iSNV richness in the multiple regression model (aOR: 1.01; 95% CI: 0.97-1.04) when controlling for Ct value.

### A signal of transmission linkage in within-host diversity

We tested whether within-host SARS-CoV-2 diversity could be used to identify transmission linkages using household membership as a proxy for probable epidemiological linkage. Pairs of individuals in the same household shared significantly more iSNVs (at a 0.2% minor allele frequency threshold, mean: 8.87 iSNVs; 95% CI: 7.81-9.92) than did pairs in different households infected with the same viral clade (mean: 2.52 iSNVs; 95% CI: 2.32-2.73) or pairs in different households infected with a different viral clade (mean: 2.92; 95% CI: 2.86-3.00), when including samples sequenced in different batches, before filtering.

Applying different variant filtering approaches dramatically reduced the number of observed iSNVs within individual samples and shared between sample pairs (Fig. S4) but did not eliminate the signal of greater levels of shared within-host diversity among household pairs than among epidemiologically unlinked pairs. After applying all filters, pairs of individuals in the same household shared significantly more iSNVs (at a 0.2% minor allele frequency threshold, mean: 0.96 iSNVs; 95% CI: 0.78-1.15) than do pairs in different households infected with the same viral clade (mean: 0.20 iSNVs; 95% CI: 0.17-0.23) or pairs in different households infected with a different viral clade (mean: 0.25; 95% CI: 0.24-0.26) (Fig. 3a) and including only samples sequenced in different batches.

Applying an increasingly stringent minor allele frequency threshold greatly reduces the number of iSNVs observed within a sample and therefore the number of iSNVs shared between samples, yet household members share more iSNVs than do epidemiologically unlinked participants. Applying a minor allele frequency threshold of 1%, for example, a mean of 0.50 iSNVs (95% CI: 0.41-0.59) are shared between pairs of samples collected from the same household and 0.073 iSNVs (95% CI: 0.060-0.086) unfiltered iSNVs are shared between pairs in different households infected with the same viral clade.

We hypothesized that minor allele frequencies of shared variants would contribute additional information about transmission beyond the number of shared variant positions; we therefore measured shared population-level diversity as the geometric mean of the sum of within-host minor allele frequencies for shared iSNVs, which we refer to below as population diversity (Methods). Across all minor allele frequency thresholds, pairs of individuals in the same household share significantly more population diversity (at a 0.2% minor allele frequency threshold, after filtering, mean: 0.028; 95% CI: 0.021-0.034) than do pairs in different households infected with the same viral clade (mean: 0.0031; 95% CI: 0.0025-0.0036) and different viral clades (mean: 0.0029; 95% CI: 0.0027-0.0030) (Fig. 3b).

In a generalized linear model for shared within-host diversity, household membership was associated with an increased odds of shared iSNVs (aOR:19.8; 95% CI: 6.39-61.40) compared to sample pairs within the same clade, after controlling for genetic distance between consensus sequences, consistent with a previous study that found household membership is the strongest predictor of shared iSNVs[9]. Longitudinal samples from an individual were also associated with an increased odds of shared iSNVs (aOR: 60.5; 95% CI:19.9-184) as were sequencing replicates (aOR: 132; 95% CI: 42.5-411).

After excluding pairs sequenced in the same batch and multiple comparisons between participants, our sample size was small (23 unique household pairs). In a generalized linear model, the number of shared iSNVs was not significantly associated with an increased odds of household membership (aOR: 1.31; 95% CI: 0.87-1.71), while a closely related consensus sequence (within 0-1 SNPs) was significantly associated with household membership (aOR: 30.38; 95% CI: 10.39-129.14). However, shared diversity as measured as the standardized sum of shared minor allele frequencies between pairs was associated with an increased odds of household membership (aOR: 1.20; 95% CI: 1.08-1.32) when controlling for closely related consensus sequence.

### Replication in a Wisconsin study

We tested the replicability of our findings in an independent study conducted in Wisconsin where SARS-CoV-2 was deep sequenced from 133 acutely-infected individuals, including members of 19 households[9]. At a frequency threshold of 0.5%, we found a similar signal that pairs of individuals in the same household shared significantly more iSNVs (mean: 9.52 iSNVs; 95% CI 8.14-10.89) than did pairs in different households infected with the same viral clade (mean: 4.28 iSNVs; 95% CI: 4.19-4.37) or pairs in different households infected with a different viral clade (mean: 1.42; 95% CI: 1.38-1.47) (Fig. 3a), in variants in filtered VCF files made publicly available from the earlier study[9] (Fig. S6; Methods). Our findings were consistent across minor allele frequency thresholds, though as in the California data, a signal of household membership was strongest when using minor allele frequency thresholds of ≤1% (Fig. S6). We found a similar signal when measuring shared population diversity as the sum of shared minor allele frequencies (Fig. S7). However, household pairs did not share significantly more diversity than epidemiologically unrelated pairs when applying all filters and a minor allele frequency threshold ≥3% (Fig. S7).

## Discussion

While most SARS-CoV-2 genomic studies focus on consensus sequences, consensus sequences may not provide the resolution needed to reconstruct transmission linkages and identify potential sources of transmission in outbreak settings, where many cases may be closely genetically related. Here, we report that within-host SARS-CoV-2 genomic variation may contribute information about transmission that may augment the information contained in viral consensus sequences. We focused on household membership as a proxy for epidemiological linkage. However, the potential utility of within-host variation would be for population surveillance or outbreak investigation, such as in hospitals or prisons, where transmission linkages are not known *a priori*.

Our measures of within-host SARS-CoV-2 diversity are consistent with those measured in previous studies when applying similar thresholds: a mean of three (range 0-5) iSNVs at a minor allele frequency ≥2% were identified in an outbreak on a fishing boat[8] and a mean of three iSNVs were reported in individuals sampled in a household study in Wisconsin above a 3% minor allele frequency threshold and consistent across sequencing replicates[9]. Further, our finding that iSNVs can be shared between epidemiologically linked individuals is consistent with previous reports that household membership is the most significant predictor of shared within-host variation[9]. Overall, SARS-CoV-2 within-host diversity is lower than that identified in other viral pathogens and, as previously reported, we find that within-host viral diversity is frequently lost during transmission[9].

As others have reported[9,23,33,34], excluding sources of noise from within-host pathogen genomic data remains a major challenge. We sequenced artificial strain mixtures of two SARS-CoV-2 variants of concern and found significant tradeoffs between sensitivity and specificity in recovery of true within-host variants as increasingly strict variant filters were applied. Applying strict minor allele frequency thresholds excludes much potential within-host variation. Additionally, in our empirical sequencing data, we find that the signal of shared within-host variation across transmission pairs is strongest when including iSNVs at low minor allele frequency thresholds.

The optimal variant identification approach may differ across applications—for example, measurements of transmission bottleneck are highly sensitive to allele frequency threshold[9,36] and may prioritize specificity, while studies of transmission might prioritize sensitivity to identify potential transmission linkages. However, again as others have highlighted, our findings underscore the need to control for other potential explanations for shared iSNVs while still prioritizing sensitivity (Box 2). Our findings suggest that for transmission inference, privileging sensitivity in variant identification may greatly improve sensitivity for recovering within-host variation, at a small cost of false positive variant calls.

Our study has several limitations. First, we focused on a convenience sample of residual samples with accompanying household information collected in California from March 2020 through May 2021. Replicating these findings in other settings and with more recently emerged SARS-CoV-2 lineages is critical to understand the generalizability of our findings. Second, our study focused on the potential epidemiological value of within-host viral variation. Our focus was on transmission linkage rather than in viral evolutionary dynamics or transmission bottlenecks, which might have different optimal variant identification approaches. Third, many groups have hypothesized that evolution within immune-compromised or immune-suppressed populations may be an important driver of the emergence of new variants of concern or interest[37–41]. Our sample collection did not enable us to test these hypotheses. Forth, the epidemiological utility of within-host variation depends on SARS-CoV-2 sampling and sequencing. Routine sequencing may always not generate sufficient depth to accurately recover within-host variation.

In conclusion, we find that SARS-CoV-2 variation within individual hosts may be shared across transmission pairs and may contribute information on transmission linkage on a backdrop of limited diversity among consensus sequences. More broadly, pathogen diversity within individual infections holds largely untapped information that may enhance the resolution of transmission inferences.

**Box 1. Determinants of within-host SARS-CoV-2 diversity**

Potential contributors to recovered SARS-CoV-2 diversity include biological determinants in addition to sampling methods.

- Biological determinants:
  - True viral population diversity, including diversity present in the infecting inoculum and diversity generated through both neutral and selective within-host processes, which in turn may be driven by the host environment, host immune status and immune history (including natural and vaccine-acquired immunity), and viral genotype.
  - Viral population size within an individual, reflecting individual infection dynamics.
- Study design:
  - Viral sampling technique and physical site of sampling may vary across studies.
  - Sequencing approach including amplicon-based, metagenomic sequencing, or other pathogen enrichment steps and sequencing platform often vary across studies.
  - Sequencing depth of coverage.
  - Prospective household transmission studies may enable infections to be identified and sampled earlier compared to samples collected through passive surveillance.
- Bioinformatic choices:
  - Read filtering, mapping and variant identification algorithms vary in sensitivity and specificity.
  - Some previous studies have required iSNVs to be identified in technical replicates[9,34].
  - Minor allele frequency thresholds vary across studies, with previous studies applying filters ranging from 2-6%[9,42].

**Box 2. Potential explanations for shared iSNVs**

As with the SARS-CoV-2 diversity present within individuals, observed shared within-host diversity could be attributable to a biological signal or the observation process.

- True positive: Transmission of a diverse infecting inoculum.
  - Within-host viral diversity can be structured temporally[33,38,41] or spatially or both.Transmitted diversity is a subset of diversity generated by within-host evolutionary processes.

- False positive:
  - Convergent or homoplastic iSNVs reflecting highly mutable sites along the genome or sites under selection.
  - Sequencing batch effects due to contamination or adapter switching during a sequencing run.
  - Artefacts of common sampling approach reflecting contamination due to similar sampling or processing environment.
  - Bioinformatic errors falling in consistent genomic regions that are difficult to map and/or identify variants.

## Data Availability

Sequence data is available at SRA (BioProject ID: PRJNA842503).

## Footnotes

### Meetings where the information has previously been presented

This information has previously presented to the California Department of Public Health COVIDNet Expert Panel.

## References

1. Turakhia Y, Thornlow B, Hinrichs AS, et al. Ultrafast Sample placement on Existing tRees (UShER) enables real-time phylogenetics for the SARS-CoV-2 pandemic. Nat Genet [Internet]. [cited 2021 Jun 3];. Available from: https://doi.org/10.1038/s41588-021-00862-7

2. Lemey P, Hong SL, Hill V, et al. Accommodating individual travel history and unsampled diversity in Bayesian phylogeographic inference of SARS-CoV-2. Nat Commun 2020 111 [Internet]. Nature Publishing Group; 2020 [cited 2022 Apr 19]; 11(1):1–14. Available from: https://www.nature.com/articles/s41467-020-18877-9

3. Korber B, Fischer WM, Gnanakaran S, et al. Tracking changes in SARS-CoV-2 Spike: evidence that D614G increases infectivity of the COVID-19 virus. Cell [Internet]. Cell Press; 2020 [cited 2020 Jul 17];. Available from: https://doi.org/10.1016/j.cell.2020.06.043.

4. Duchene S, Featherstone L, Haritopoulou-Sinanidou M, Rambaut A, Lemey P, Baele G. Temporal signal and the phylodynamic threshold of SARS-CoV-2. Virus Evol [Internet]. Oxford Academic; 2020 [cited 2022 Mar 14]; 6(2). Available from: https://academic.oup.com/ve/article/6/2/veaa061/5894560

5. Borges V, Isidro J, Macedo F, et al. Nosocomial Outbreak of SARS-CoV-2 in a “Non-COVID-19” Hospital Ward: Virus Genome Sequencing as a Key Tool to Understand Cryptic Transmission. Viruses. MDPI AG; 2021; 13(4).

6. Choi EM, Chu DKW, Cheng PKC, et al. In-Flight Transmission of SARS-CoV-2. Emerg Infect Dis [Internet]. Centers for Disease Control and Prevention; 2020 [cited 2022 May 23]; 26(11):2713. Available from: /pmc/articles/PMC7588512/

7. Lemieux, Lemieux JE, Siddle KJ, et al. Phylogenetic analysis of SARS-CoV-2 in Boston highlights the impact of superspreading events. Science (80-) [Internet]. 2020 [cited 2020 Dec 16]; :eabe3261. Available from: https://www.sciencemag.org/lookup/doi/10.1126/science.abe3261

8. Hannon WW, Roychoudhury P, Xie H, et al. Narrow transmission bottlenecks and limited within-host viral diversity during a SARS-CoV-2 outbreak on a fishing boat. bioRxiv [Internet]. Cold Spring Harbor Laboratory; 2022 [cited 2022 Feb 16]; :2022.02.09.479546. Available from: https://www.biorxiv.org/content/10.1101/2022.02.09.479546v1

9. Braun KM, Moreno GK, Wagner C, et al. Acute SARS-CoV-2 infections harbor limited within-host diversity and transmit via tight transmission bottlenecks. PLoS Pathog [Internet]. Public Library of Science; 2021 [cited 2022 Feb 10]; 17(8):e1009849. Available from: https://journals.plos.org/plospathogens/article?id=10.1371/journal.ppat.1009849

10. Meredith LW, Hamilton WL, Warne B, et al. Rapid implementation of SARS-CoV-2 sequencing to investigate cases of health-care associated COVID-19: a prospective genomic surveillance study. Lancet Infect Dis [Internet]. 2020 [cited 2020 Jul 17]; 20(11):1263–1272. Available from: www.thelancet.com/infectionPublishedonline

11. Tonkin-Hill G, Ling C, Chaguza C, et al. Pneumococcal within-host diversity during colonisation, transmission and treatment. [cited 2022 Mar 7];. Available from: https://doi.org/10.1101/2022.02.20.480002

12. Wymant C, Hall M, Ratmann O, et al. PHYLOSCANNER: Inferring transmission from within-and between-host pathogen genetic diversity. Mol Biol Evol [Internet]. Oxford University Press; 2018 [cited 2020 Oct 1]; 35(3):719–733. Available from: http://creativecommons.

13. Leitner T. Phylogenetics in HIV transmission: Taking within-host diversity into account. Curr Opin HIV AIDS [Internet]. Lippincott Williams and Wilkins; 2019 [cited 2021 Jul 16]; 14(3):181– 187. Available from: https://journals.lww.com/co-hivandaids/Fulltext/2019/05000/Phylogenetics_in_HIV_transmission__taking.6.aspx

14. Martin MA, Koelle K. Comment on “Genomic epidemiology of superspreading events in Austria reveals mutational dynamics and transmission properties of SARS-CoV-2.” Sci Transl Med [Internet]. American Association for the Advancement of Science; 2021 [cited 2021 Nov 10]; 13(617):1803. Available from: https://www.science.org/doi/abs/10.1126/scitranslmed.abh1803

15. San JE, Ngcapu S, Kanzi AM, et al. Transmission dynamics of SARS-CoV-2 within-host diversity in two major hospital outbreaks in South Africa. Virus Evol [Internet]. Oxford Academic; 2021 [cited 2022 May 23]; 7(1):41. Available from: https://academic.oup.com/ve/article/7/1/veab041/6248115

16. Altamirano J, Govindarajan P, Blomkalns A, et al. 401. Natural History of Shedding and Household Transmission of Severe Acute Respiratory Syndrome Coronavirus 2 Using Intensive High-Resolution Sampling. Open Forum Infect Dis. Oxford University Press (OUP); 2021; 8(Supplement_1):S302–S302.

17. Jagannathan P, Andrews JR, Bonilla H, et al. Peginterferon Lambda-1a for treatment of outpatients with uncomplicated COVID-19: a randomized placebo-controlled trial. Nat Commun [Internet]. 2021 [cited 2021 Apr 18]; 12(1). Available from: https://doi.org/10.1038/s41467-021-22177-1

18. Holubar M, Subramanian A, Purington N, et al. Favipiravir for treatment of outpatients with asymptomatic or uncomplicated COVID-19: a double-blind randomized, placebo-controlled, phase 2 trial. Clin Infect Dis [Internet]. 2022 [cited 2022 May 18];. Available from: https://academic.oup.com/cid/advance-article/doi/10.1093/cid/ciac312/6572081

19. Verma R, Kim E, Degner N, Walter KS, Singh U, Andrews JR. Variation in SARS-CoV-2 bioaerosol production in exhaled breath. Open Forum Infect Dis [Internet]. 2021 [cited 2021 Dec 11];. Available from: https://academic.oup.com/ofid/advance-article/doi/10.1093/ofid/ofab600/6447624

20. Benjamin F, Diana R, Betteridge E, et al. COVID-19 ARTIC v3 Illumina library construction and sequencing protocol V.5. Available from: https://dx.doi.org/10.17504/protocols.io.bibtkann

21. Ewels PA, Peltzer A, Fillinger S, et al. The nf-core framework for community-curated bioinformatics pipelines [Internet]. Nat. Biotechnol. Nature Research; 2020 [cited 2021 Jun 4]. p. 276–278. Available from: https://doi.org/10.1038/s41587-020-0446-y.

22. Langmead B, Salzberg SL. Fast gapped-read alignment with Bowtie 2. Nat Methods. 2012; 9(4):357–359.

23. Grubaugh ND, Gangavarapu K, Quick J, et al. An amplicon-based sequencing framework for accurately measuring intrahost virus diversity using PrimalSeq and iVar. Genome Biol [Internet]. 2019 [cited 2020 May 26]; 20(1). Available from: https://doi.org/10.1186/s13059-018-1618-7

24. Wood DE, Salzberg SL. Kraken: Ultrafast metagenomic sequence classification using exact alignments. Genome Biol [Internet]. BioMed Central; 2014 [cited 2019 Apr 1]; 15(3):R46. Available from: http://genomebiology.biomedcentral.com/articles/10.1186/gb-2014-15-3-r46

25. Li H. A statistical framework for SNP calling, mutation discovery, association mapping and population genetical parameter estimation from sequencing data. Bioinformatics [Internet]. 2011 [cited 2019 Aug 11]; 27(21):2987–2993. Available from: http://www.ncbi.nlm.nih.gov/pubmed/21903627

26. Rambaut A, Holmes EC, O’Toole Á, et al. A dynamic nomenclature proposal for SARS-CoV-2 lineages to assist genomic epidemiology. Nat Microbiol [Internet]. 2020 [cited 2021 Jun 4]; 5(11):1403–1407. Available from: https://www.nature.com/articles/s41564-020-0770-5.pdf

27. Maio N De, Walker C, Borges R, Weilguny L, g S, Goldman N. Masking strategies for SARS-CoV-2 alignments. 2020.

28. Nakamura T, Yamada KD, Tomii K, Katoh K. Parallelization of MAFFT for large-scale multiple sequence alignments. Bioinformatics [Internet]. Oxford University Press; 2018 [cited 2020 Oct 8]; 34(14):2490–2492. Available from: https://mafft.cbrc.jp/alignment/software/mpi.html.

29. Paradis E, Schliep K. Ape 5.0: An environment for modern phylogenetics and evolutionary analyses in R. Schwartz R, editor. Bioinformatics [Internet]. Narnia; 2019 [cited 2019 Apr 10]; 35(3):526–528. Available from: https://academic.oup.com/bioinformatics/article/35/3/526/5055127

30. Minh BQ, Schmidt HA, Chernomor O, et al. IQ-TREE 2: New Models and Efficient Methods for Phylogenetic Inference in the Genomic Era. Mol Biol Evol [Internet]. Oxford Academic; 2020 [cited 2022 May 17]; 37(5):1530–1534. Available from: https://academic.oup.com/mbe/article/37/5/1530/5721363

31. Bates D, Mächler M, Bolker BM, Walker SC. Fitting Linear Mixed-Effects Models Using lme4. J Stat Softw [Internet]. American Statistical Association; 2015 [cited 2022 Apr 12]; 67(1):1–48. Available from: https://www.jstatsoft.org/index.php/jss/article/view/v067i01

32. Mccrone JT, Lauring S. Measurements of Intrahost Viral Diversity Are Extremely Sensitive to Systematic Errors in Variant Calling. J Virol. 2016; 90(15):6884–6895.

33. Valesano AL, Rumfelt KE, Dimcheff DE, et al. Temporal dynamics of SARS-CoV-2 mutation accumulation within and across infected hosts. Pekosz A, editor. PLOS Pathog [Internet]. Public Library of Science; 2021 [cited 2021 Apr 16]; 17(4):e1009499. Available from: https://dx.plos.org/10.1371/journal.ppat.1009499

34. Hannon WW, Roychoudhury P, Xie H, et al. Narrow transmission bottlenecks and limited within-host viral diversity during a SARS-CoV-2 outbreak on a fishing boat. [cited 2022 Feb 16];. Available from: https://doi.org/10.1101/2022.02.09.479546

35. Lythgoe KA, Hall M, Ferretti L, et al. SARS-CoV-2 within-host diversity and transmission. Science (80-) [Internet]. American Association for the Advancement of Science (AAAS); 2021 [cited 2021 Apr 21]; 372(6539):eabg0821. Available from: https://doi.org/10.1126/science.abg0821

36. Martin MA, Koelle K. Comment on “Genomic epidemiology of superspreading events in Austria reveals mutational dynamics and transmission properties of SARS-CoV-2.” Sci Transl Med [Internet]. American Association for the Advancement of Science; 2021 [cited 2021 Oct 28]; 13(617):1803. Available from: https://www.science.org/doi/10.1126/scitranslmed.abh1803

37. Rambaut A, Loman N, Pybus O, et al. Preliminary genomic characterisation of an emergent SARS-CoV-2 lineage in the UK defined by a novel set of spike mutations - SARS-CoV-2 coronavirus / nCoV-2019 Genomic Epidemiology - Virological [Internet]. https://Virological.org. 2020 x[cited 2021 Feb 14]. Available from: https://virological.org/t/preliminary-genomic-characterisation-of-an-emergent-sars-cov-2-lineage-in-the-uk-defined-by-a-novel-set-of-spike-mutations/563

38. Kemp SA, Collier DA, Datir RP, et al. SARS-CoV-2 evolution during treatment of chronic infection. Nat 2021 5927853 [Internet]. Nature Publishing Group; 2021 [cited 2022 May 2]; 592(7853):277–282. Available from: https://www.nature.com/articles/s41586-021-03291-y

39. Weigang S, Fuchs J, Zimmer G, et al. Within-host evolution of SARS-CoV-2 in an immunosuppressed COVID-19 patient as a source of immune escape variants. Nat Commun 2021 121 [Internet]. Nature Publishing Group; 2021 [cited 2022 May 11]; 12(1):1–12. Available from: https://www.nature.com/articles/s41467-021-26602-3

40. Bessière P, Volmer R. From one to many: The within-host rise of viral variants. PLOS Pathog [Internet]. Public Library of Science; 2021 [cited 2022 May 11]; 17(9):e1009811. Available from: https://journals.plos.org/plospathogens/article?id=10.1371/journal.ppat.1009811

41. Choi B, Choudhary MC, Regan J, et al. Persistence and Evolution of SARS-CoV-2 in an Immunocompromised Host. N Engl J Med [Internet]. Massachusetts Medical Society; 2020 [cited 2021 Jan 7];. Available from: https://www.nejm.org/doi/full/10.1056/NEJMc2031364

42. Maio N De, Worby CJ, Wilson DJ, Stoesser N. Bayesian reconstruction of transmission within outbreaks using genomic variants. Koelle K, editor. PLoS Comput Biol [Internet]. Public Library of Science; 2018 [cited 2020 Oct 1]; 14(4):e1006117. Available from: https://dx.plos.org/10.1371/journal.pcbi.1006117

43. Aksamentov I, Roemer C, Hodcroft EB, Neher RA. Nextclade: clade assignment, mutation calling and quality control for viral genomes. J Open Source Softw [Internet]. The Open Journal; 2021 [cited 2022 Apr 11]; 6(67):3773. Available from: https://joss.theoj.org/papers/10.21105/joss.03773

44. Ewels PA, Peltzer A, Fillinger S, et al. The nf-core framework for community-curated bioinformatics pipelines. Nat Biotechnol 2020 383 [Internet]. Nature Publishing Group; 2020 [cited 2022 Apr 11]; 38(3):276–278. Available from: https://www.nature.com/articles/s41587-020-0439-x

