## Supplementary Information for "Shared within-host SARS-CoV-2 variation in households"

**Supplementary Methods**

**Serial dilution experiment.**

We conducted a serial dilution experiment, mixing synthetic RNA controls (Twist Biosciences) representing the Alpha (B.1.1.7) and Beta (B.1.351) variants so that the minor variant (Beta) comprised 0, 0.25, 0.5, 2, 5, 10, and 100% of the mixture. We then serially diluted mixtures so that samples had a total of 10^1^-10^5^ total RNA copies (Fig. 1). Consensus sequences of the Alpha and Beta variant synthetic controls differ at 43 sites. We deep sequenced artificial synthetic mixtures using the ARTIC v3 tiled amplicon sequencing protocol[1], identified iSNVs with the *viralrecon* pipeline[2], and determined the sensitivity and specificity of our variant calling pipeline to recovering true variation within our synthetic mixtures.

We created a “truth VCF” of variant positions expected in our strain mixtures with respect to the Wuhan-1 reference genome by aligning EPI_ISL_710528 (Alpha variant) and the EPI_ISL_678597 (Beta variant) reference genomes to the MN908947.3 reference genome (MN908947.3) with *mafft*[3] and *snp-sites*[4]. Consensus sequences of the Alpha and Beta controls differed at 43 sites.

We then determined the sensitivity and specificity of our variant calling pipeline to recovering true variation within our synthetic mixtures. We quantified and visualized the sensitivity, specificity, and receiver operating characteristic (ROC) curves of our pipeline in identifying true iSNVs with the R package *yardstick*[5]. We defined minor allele frequency as the alternate allele frequency or 1- the alternative allele frequency, when the alternate allele frequency was greater than 0.5. We additionally used the alternate allele call to distinguish between true positive and false positive alternative alleles. We stratified ROC curves by the proportion of minor variant in the input strain mixture and by dilution. We determined Pearson’s correlation coefficients for the observed and expected minor allele frequencies, stratified by dilution with the R package *stats*[6]*.*

**Sample RT-qPCR testing**

We used CDC-qualified primers and TaqMan probes to amplify SARS-CoV-2 nucleocapsid (N) gene (Supplementary Methods). Human RNaseP was used as a quality control [16]. Briefly, TaqPath 1-step RT-qPCR mastermix (Invitrogen, Darmstadt, Germany) was used in a 20-µL reaction volume, and the samples were analyzed on a StepOne-Plus (Applied Biosystems) instrument using the following program: 10 minutes at 50ºC for reverse transcription, followed by 3 minutes at 95ºC and 40 cycles of 10 seconds at 95ºC, 15 seconds at 56ºC, and 5 seconds at 72ºC. We estimated viral RNA copies/sample from a standard curve using a pET21b+ plasmid (GenScript) with the N-gene. The cycle threshold (Ct) cutoff for positive samples was <38.

**Library preparation and sequencing**

We followed the ARTIC v3 Illumina library preparation and sequencing protocols[1] and sequenced amplicons on an Illumina MiSeq platform. Briefly, 10μL template RNA was reverse transcribed to cDNA using Lunascript RT supermix (New England Biolabs). 1X Q5 Hotstart mastermix (New England Biolabs) was used to amplify cDNA using each of the two ARTIC v3 primer pools A and B (17). The amplicons were purified and pooled for dA-tail repair with NEBNext Ultra II End prep enzyme mix, followed by adapter ligation (NEBNext Ultra II ligation mix and 15uM NEBNext Adaptor). Dual indexing was performed using i5 and i7 adapters (New England Biolabs) as per the manufacturer’s instructions using 2X KAPA HiFi HotStart mastermix (Kapa Biosystems). The pooled samples were analyzed on Illumina MiSeq using V2 reagent kit 500 cycles (212/212 cycles).

**Phylogenetic inference**

We aligned consensus genome sequences with *mafft v7.310*[28], removing sequences with more than 20% ambiguous characters, and masked previously reported problematic sites[27] in alignments. We used the R package *ape* to measure pairwise SNP distance between consensus sequences[29]. We inferred a maximum likelihood phylogeny with IQ-TREE v 1.6.11[30] using a GTR substitution model with empirical base frequencies, a FreeRate model with two rate categories, and ascertainment bias correction for invariant sites not included in our alignment, identified by IQ-TREE as the best fit model.

**Supplementary Text**

**Sequencing artificial SARS-CoV-2 RNA mixtures reveals tradeoffs in sensitivity and specificity of within-host variant identification.**

A major challenge in studies of within-host pathogen diversity is in distinguishing true, low frequency intrahost nucleotide variants (iSNVs) from sequencing or bioinformatic errors^2^. The relative costs of false positive and false negative iSNVs may vary for different objectives. For example, false positive iSNVs—if they are shared between individuals—may upwardly bias estimates of the transmission bottleneck, or transmitted viral diversity^3^. If, alternatively, our goal is epidemiological inference, an increased sensitivity for iSNVs, may improve sensitivity in detecting shared iSNVs, and possible transmission linkages, at some expense to specificity.

To determine a minor allele frequency threshold above which we could accurately identify iSNVs, we mixed synthetic RNA controls representing the Alpha (B.1.1.7) and Beta (B.1.351) SARS-CoV-2 variants, so that the minor variant comprised 0-10% of the total RNA. To determine viral load requirements for accurate identification of iSNVs, we serially diluted samples to a total of 10^1^-10^5^ total RNA copies per uL, corresponding to Ct of 21.2 to 33.9 (Fig. 1a).

Prior to filtering, our sequencing and variant identification pipeline had an area under the curve (AUC) of greater than 0.79 for identifying true iSNVs across all minor allele frequencies present in mixtures with 10^3^ virions/uL and greater, corresponding to RT-qPCR Ct values of 27.6 and lower (Fig. 1b). Variant recovery was consistent across minor allele frequencies: for mixtures with 10^3^ viral genomes per uL, AUCs for all minor allele frequencies were 0.79 (Fig. 1b) prior to filtering. Receiver operating characteristic (ROC) curves across dilutions have sharp inflection points, indicating that decreases in the minor allele frequency threshold rapidly lead to strong improvements in sensitivity at a negligible cost to specificity (Fig. 1b).

Before filtering, observed minor allele frequencies were closely correlated with expected minor allele frequencies across all mixtures in which the Beta variant was present at a frequency as low as 0.25% (Fig. 1c), with a correlation coefficient exceeding 0.958 for true iSNV positions in mixtures of 10^3^ virions/uL and greater, corresponding to RT-qPCR Ct values of 27.6 and lower. The majority, 90.1% (247/274) of false positive iSNVs were at frequencies of less than 0.2% (Fig. 1c). False positive iSNVs were frequently repeated across artificial mixtures. Among the 58 unique false positive positions, 67.2% (39/58) were repeated, and 10.3% (6/58) positions were repeated across 25% (10/40) artificial mixture samples (Fig. 1c).

To test whether commonly applied filters would improve overall accuracy, we applied three variant filters: a filter for iSNV quality from iVar^5^, a quality score filter (Phred score >40), a depth filter (of both major and minor alleles > 5X), and all filters. Filters generally slightly decreased AUC compared to unfiltered variants (Table S1), because of excluding true positive iSNVs; for example, the AUC for mixtures with 10^3^ virions decreased from 0.79 to 0.78 after applying all filters.

We additionally tested the effect of filtering by minor allele frequency. The underlying true distribution of minor allele frequencies in individual infections is unknown, making it difficult to measure the true sensitivity and specificity of iSNV recovery. We therefore examined possible tradeoffs in sensitivity and specificity across a range of minor allele frequencies (Fig. S1a, Table S2). As expected, when the true minor allele frequency is 10%, sensitivity and specificity of recovering true minor variants remain high after filtering. However, when minor alleles are present at low frequencies, imposing a minor allele frequency threshold rapidly excludes true variation and sensitivity declines to 0. In contrast, there is a minor cost to specificity when including low frequency iSNVs, after the iSNVs below 0.2% are excluded (Fig. S1b).

**Figure S1. Sensitivity (a) and specificity (b) of recovering true iSNVs distinguishing the Alpha and Beta variants in serial mixture experiments across different minor allele frequency thresholds**, or thresholds above which an iSNV is included. Colors indicate the expected minor allele frequency, the proportion of the minor variant included in the full strain mixture.

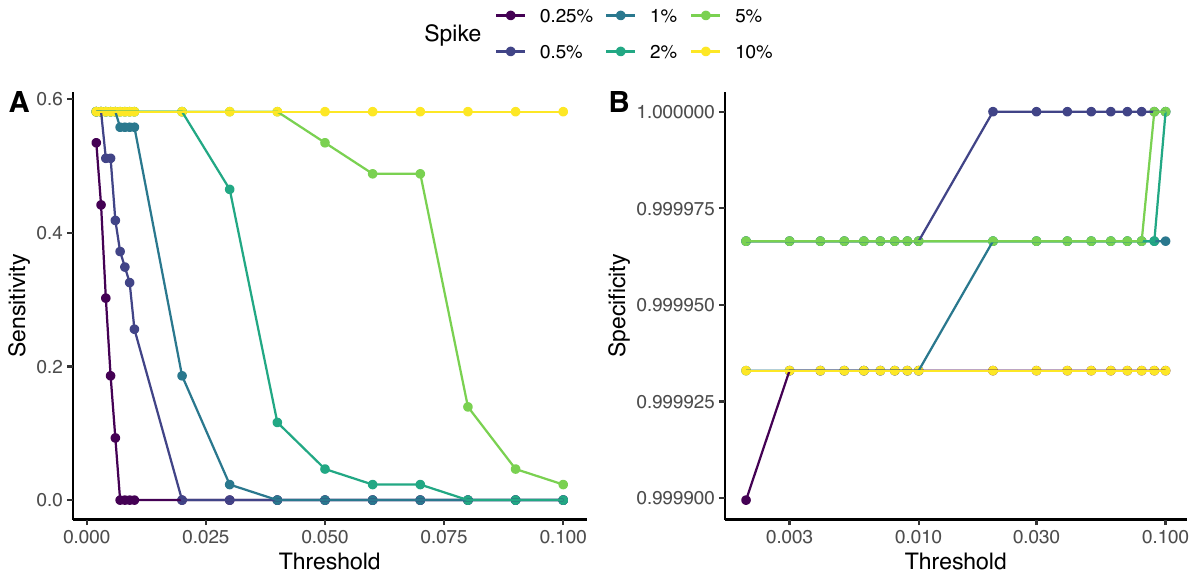

**Figure S2. iSNVs identified within individual California samples and pairwise shared iSNVs across samples.** From our genomic collection from California, pairwise comparisons of the number of shared iSNVs, defined as a shared minor allele present at the same genomic position, identified across different minor allele frequency thresholds, after applying all filters. Points and error bars indicate mean and 95% confidence intervals and are colored by comparison type. For confidence intervals spanning, 0, we set lower limits to 10^-4^. Each pair is assigned to a unique category. Sample: iSNVs identified within a single sample; Sequencing replicate: pairs of samples from the same individual and same time point; Within-host: pairs of samples from the same individual collected on different days; Household: pairs of individuals from the same household; Clade: pairs of individuals outside households infected with the same Nextclade clade; and Outside clade: pairs of individuals outside households infected with different Nextclade clades. Pairwise comparisons include only samples sequenced on different sequencing batches.

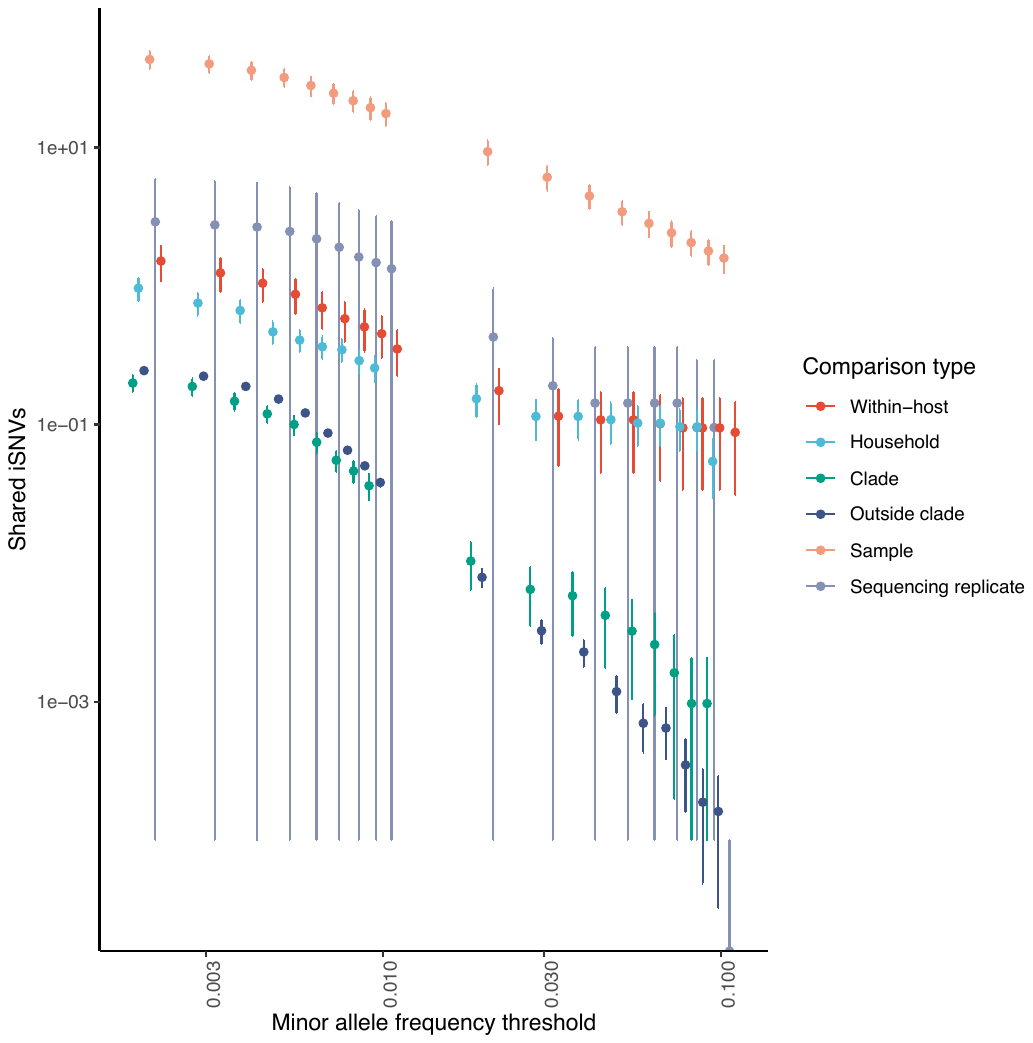

**Figure S3. Pairwise shared iSNVs among California samples with closely related consensus sequences.** To test if shared iSNVs provided information to augment that in consensus sequences, we examined only pairwise comparisons of sequences with consensus sequences differing by 0-1 SNP, identified across different minor allele frequency thresholds, after applying all filters. Points and error bars indicate mean and 95% confidence intervals and are colored by comparison type. Each pair is assigned to a unique category. Within-host: pairs of samples from the same individual collected on different days; Household: pairs of individuals from the same household; Clade: pairs of individuals outside households infected with the same Nextclade clade; and Outside clade: pairs of individuals outside households infected with different Nextclade clades. Pairwise comparisons include only samples sequenced on different sequencing batches.

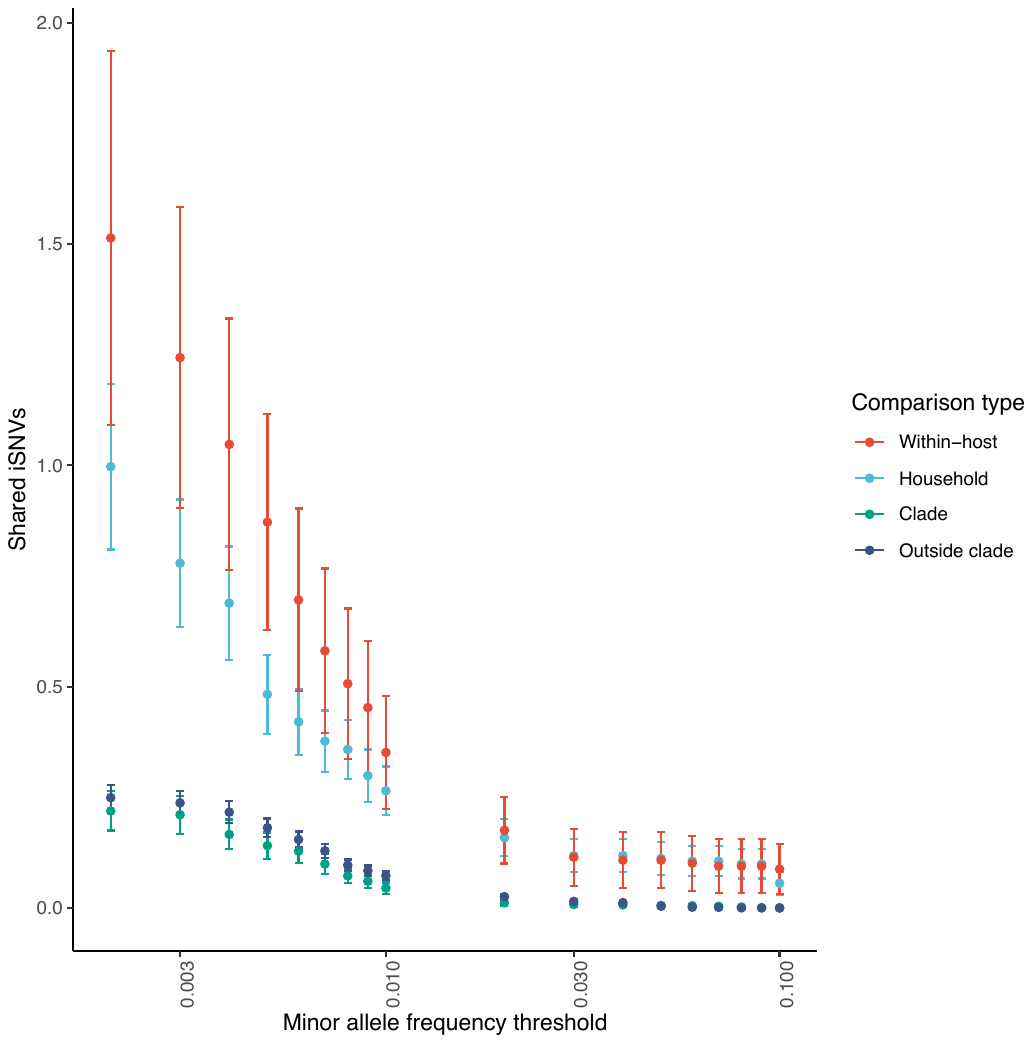

**Figure S4. Pairwise shared iSNVs among California samples across sequencing batches and variant filters.** For our genomic collection from California, pairwise comparisons of the number of shared iSNVs identified across different minor allele frequency thresholds and variant filters. Facets indicate sequencing batch. Batch: samples sequenced on the same Illumina sequencing run; Different batch: samples sequenced on different Illumina runs. Plots indicate variant identification filters applied. Unfiltered: no variant filtering; Frequent FP filter: positions of false positive iSNVs identified in more than one sample in the serial dilution experiment are excluded; iVar filter: positions failing the iVar filter for iSNV quality are excluded^4^; Quality filter: quality score filter (Phred score >40); Depth filter: a depth filter (of both major and minor alleles > 5X); and All filters. Points and error bars indicate mean and 95% confidence intervals and are colored by comparison type. Each pair is assigned to a unique category. Within-host: pairs of samples from the same individual collected on different days; Household: pairs of individuals from the same household; Clade: pairs of individuals outside households infected with the same Nextclade clade; and Outside clade: pairs of individuals outside households infected with different Nextclade clades.

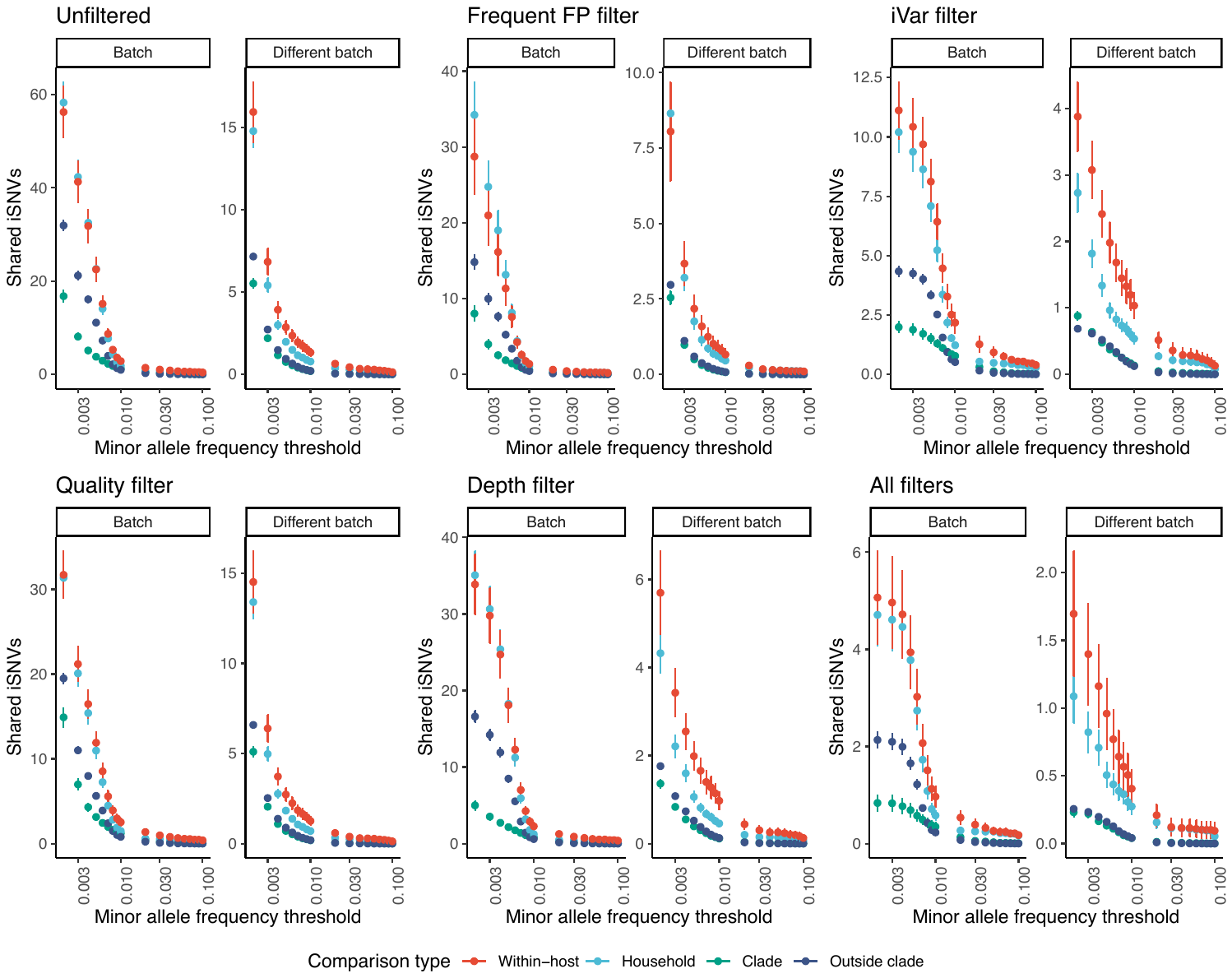

**Figure S5. Pairwise shared within-host population diversity among California samples across sequencing batches and variant filters.** For our genomic collection from California, pairwise comparisons of shared population diversity, as measured by the geometric mean of the sum of within-host minor allele frequencies for shared iSNVs, identified across different minor allele frequency thresholds and variant filters. Facets indicate sequencing batch. Batch: samples sequenced on the same Illumina sequencing run; Different batch: samples sequenced on different Illumina runs. Plots indicate variant identification filters applied. Unfiltered: no variant filtering; Frequent FP filter: positions of false positive iSNVs identified in more than one sample in the serial dilution experiment are excluded; iVar filter: positions failing the iVar filter for iSNV quality are excluded^4^; Quality filter: quality score filter (Phred score >40); Depth filter: a depth filter (of both major and minor alleles > 5X); and All filters. Points and error bars indicate mean and 95% confidence intervals and are colored by comparison type. Each pair is assigned to a unique category. Within-host: pairs of samples from the same individual collected on different days; Household: pairs of individuals from the same household; Clade: pairs of individuals outside households infected with the same Nextclade clade; and Outside clade: pairs of individuals outside households infected with different Nextclade clades.

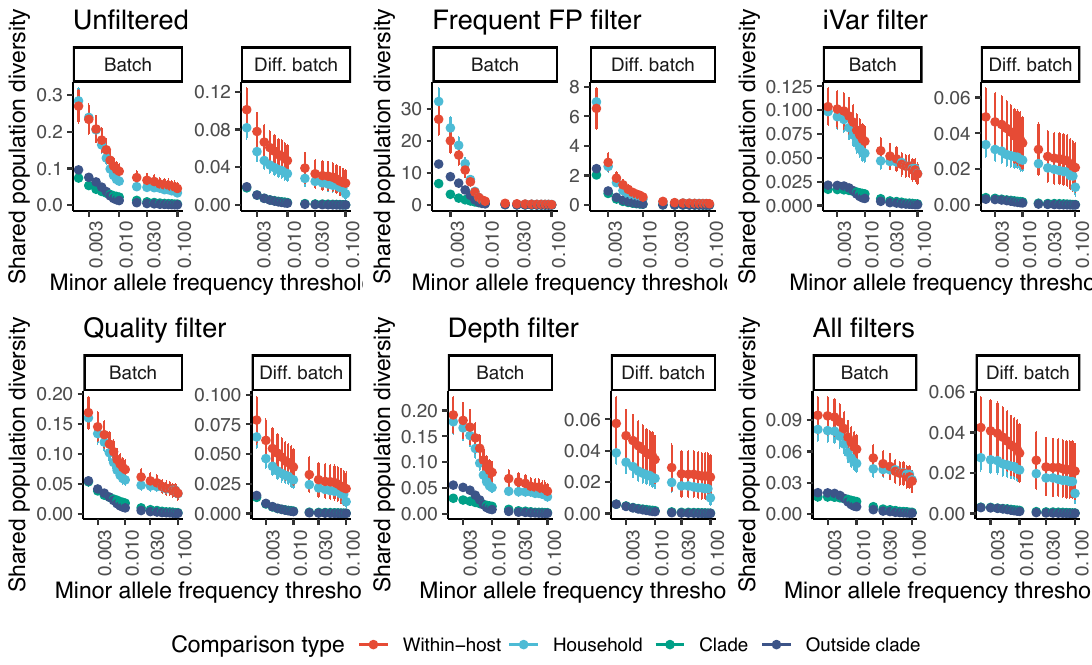

**Figure S6. Pairwise shared iSNVs in previously published variants identified in Wisconsin.** To test if our findings were robust to variant identification pipeline, we also investigated shared iSNVs in variants identified within a genomic epidemiology study in Wisconsin^8^. Pairwise comparisons of the number of shared within-host iSNVs, defined as a shared minor allele present at the same genomic position, identified across different minor allele frequency thresholds. Points and error bars indicate mean and 95% confidence intervals and are colored by comparison type. Each pair is assigned to a unique category. Household: pairs of individuals from the same households with consensus sequences within 0-2 SNPs; Clade: pairs of individuals outside households infected with the same Nextclade clade; and Outside clade: pairs of individuals outside households infected with different Nextclade clades. As in the previous study, we excluded household pairs from comparisons with consensus genomes that differed by more than 2 SNPs^8^.

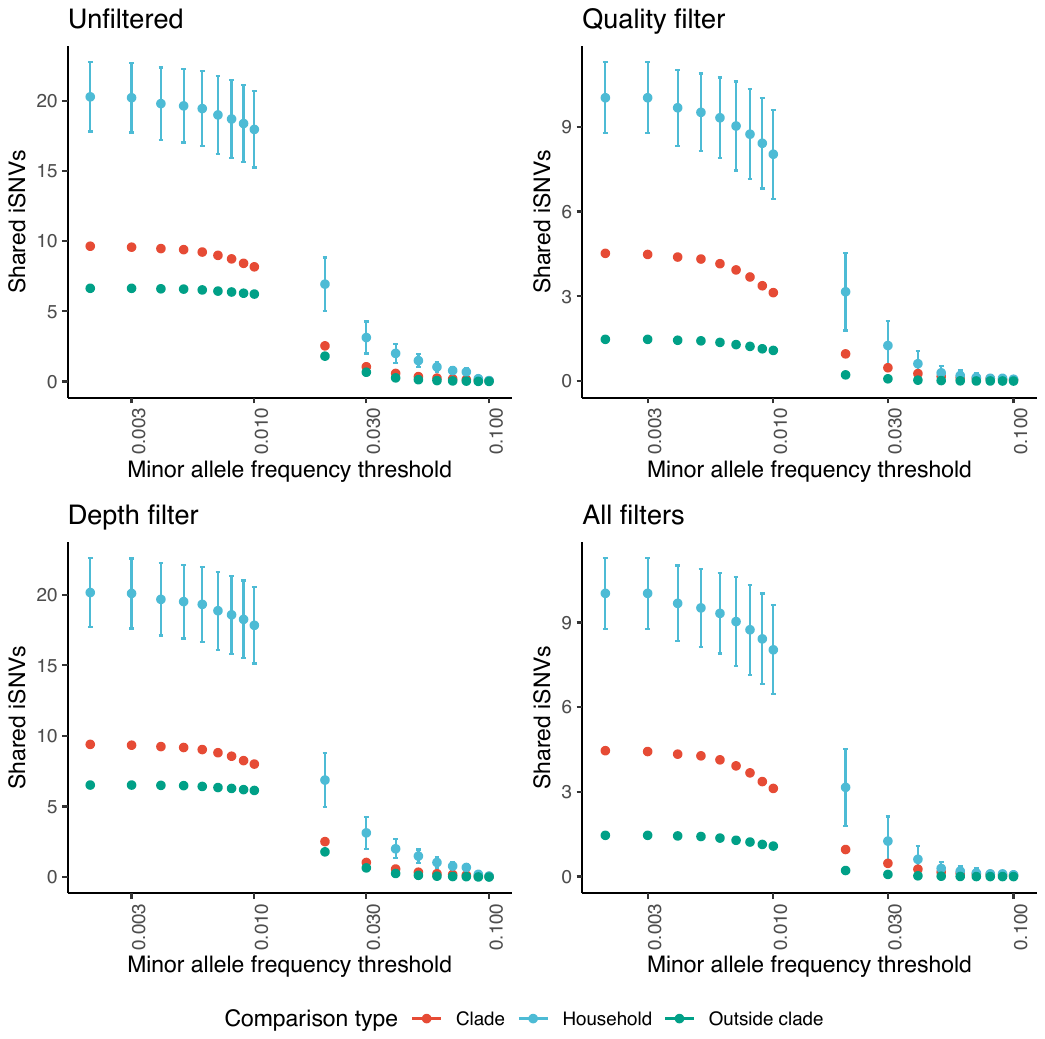

**Figure S7. Pairwise shared SARS-Cov-2 population diversity in previously published variants identified in Wisconsin.** To confirm that our findings were robust to variant identification pipeline, we also investigated shared iSNVs in variants identified within a genomic epidemiology study in Wisconsin^8^. Pairwise comparisons of the number of shared within-host iSNVs, defined as a shared minor allele present at the same genomic position, identified across different minor allele frequency thresholds. Points and error bars indicate mean and 95% confidence intervals and are colored by comparison type. Each pair is assigned to a unique category. Household: pairs of individuals from the same households with consensus sequences within 0-2 SNPs; Clade: pairs of individuals outside households infected with the same Nextclade clade; and Outside clade: pairs of individuals outside households infected with different Nextclade clades. As in the previous study, we excluded household pairs from comparisons with consensus genomes that differed by more than 2 SNPs^8^.

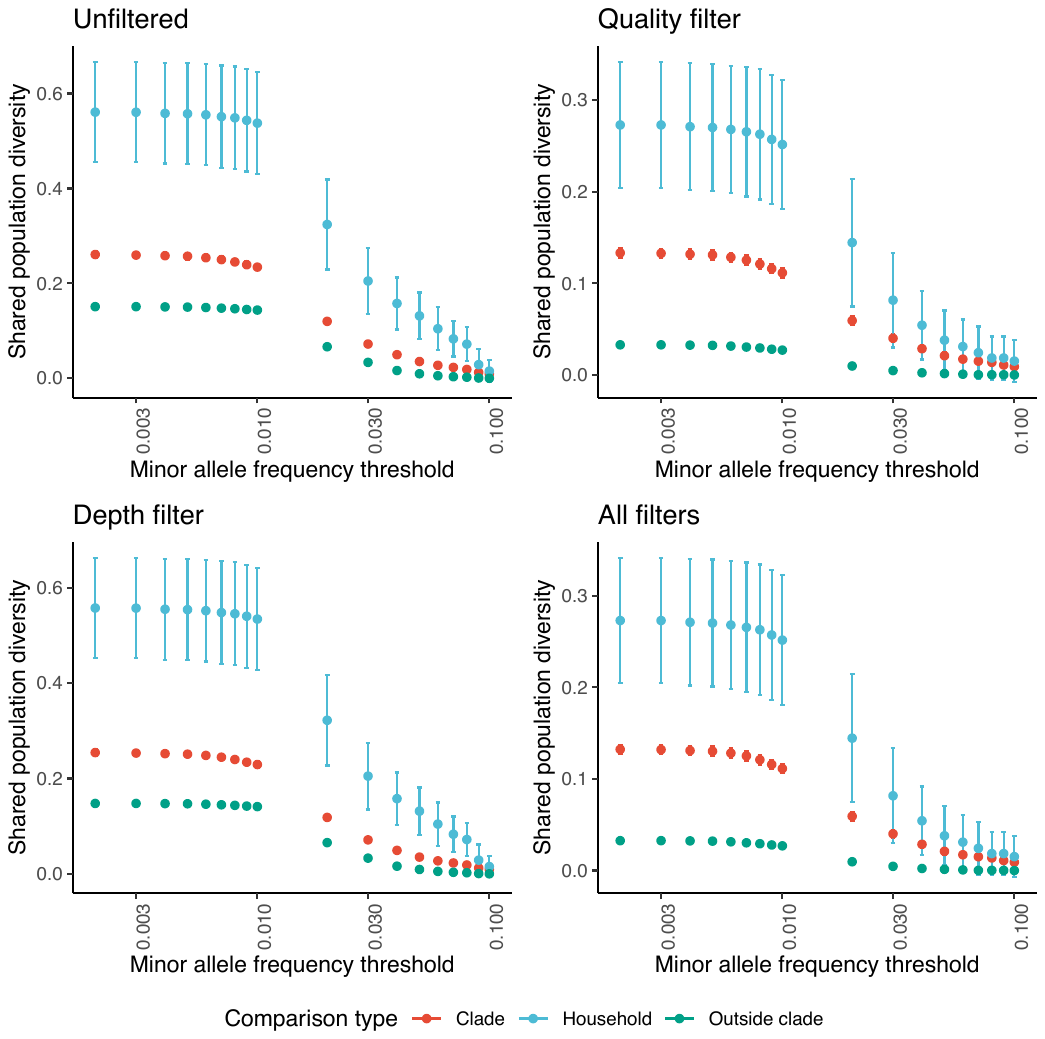

**Table S1.** **Area under the curve (AUC) for detecting true minor alleles in an artificial strain mixture experiment.** AUC for detecting minor alleles among all samples, stratified by viral dilution, total viral copies per uL. Columns indicate variant calling filter applied. None: unfiltered; iVar: positions failing the iVar filter for iSNV quality are excluded^4^; Qual: quality score filter (Phred score >40); Depth: a depth filter (of both major and minor alleles > 5X); and All filters.

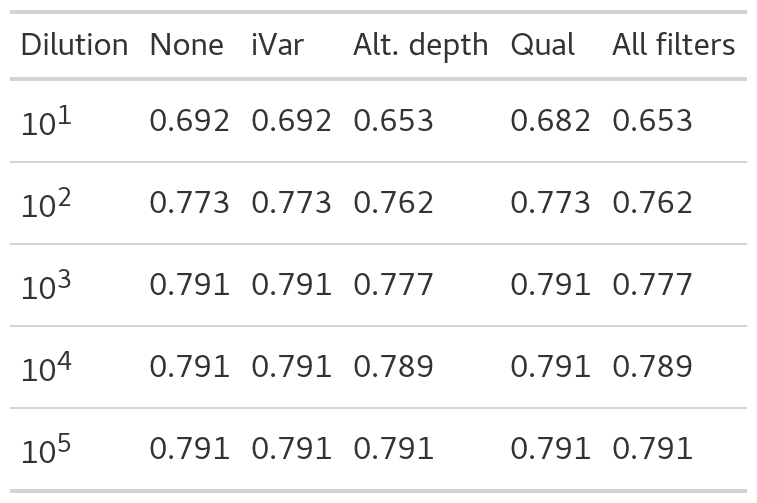

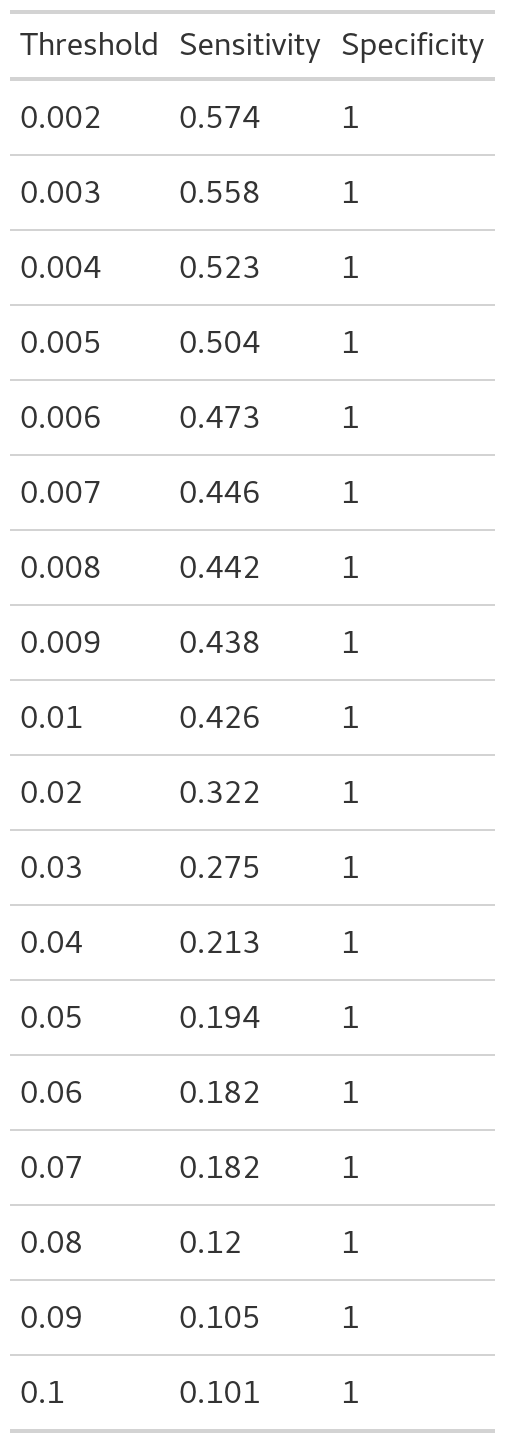
**Table S2.** **Sensitivity and specificity in detecting true iSNVs in an artificial strain mixtures experiment across minor allele frequency thresholds.** We report the sensitivity and specificity for iSNV detection after filtering at different minor allele frequency thresholds for samples diluted to10^3^ viral genomes/uL. Each row includes samples in which the Beta variant comprises 0.25-10% of the total viral RNA input.

**Table S3.** **Sensitivity and specificity in detecting true iSNVs in an artificial strain mixtures experiment across true minor allele frequencies.** We report the sensitivity and specificity for iSNV detection across samples in which the Beta variant comprises 0.25%-10% of the total viral RNA. We report sensitivity and specificity for variants were filtered at a minor allele frequency threshold of 0.5% and for samples diluted to10^3^ viral genomes/uL.

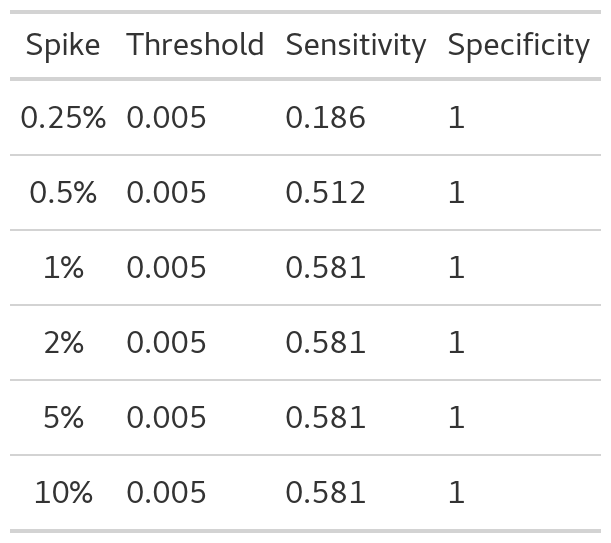
